# T cell responses to SARS-CoV-2 vaccination in people with multiple sclerosis differ between disease-modifying therapies

**DOI:** 10.1101/2022.08.25.22279202

**Authors:** Asia-Sophia Wolf, Anthony Ravussin, Marton König, Mathias H. Øverås, Guri Solum, Ingrid Fadum Kjønstad, Adity Chopra, Trygve Holmøy, Hanne F. Harbo, Silje Watterdal Syversen, Kristin Kaasen Jørgensen, Einar August Høgestøl, Jon Torgils Vaage, Elisabeth G. Celius, Fridtjof Lund-Johansen, Ludvig A. Munthe, Gro Owren Nygaard, Siri Mjaaland

## Abstract

Immune responses in people with multiple sclerosis (pwMS) on disease-modifying therapies (DMTs) have been of significant interest throughout the COVID-19 pandemic. Lymphocyte-targeting immunotherapies including anti-CD20 treatments and sphingosine-1-phosphate receptor (S1PR) modulators attenuate antibody responses after vaccination. Evaluation of cellular responses after vaccination is therefore of particular importance in these populations. In this study, we analysed CD4 and CD8 T cell functional responses to SARS-CoV-2 spike peptides in healthy controls and pwMS on five different DMTs by flow cytometry. Although pwMS on anti-CD20 and S1PR therapies had low antibody responses after both 2 and 3 vaccine doses, T cell responses in pwMS on anti-CD20 therapies were preserved after a third vaccination, even when additional anti-CD20 treatment was administered between vaccine doses 2 and 3. PwMS taking S1PR modulators had low detectable T cell responses in peripheral blood. CD4 and CD8 T cell responses to SARS-CoV-2 variants of concern Delta and Omicron were lower than to the ancestral Wuhan-Hu-1 variant. Our results indicate the importance of assessing both cellular and humoral responses after vaccination and suggest that even in the absence of robust antibody responses vaccination can generate immune responses in pwMS.

## Introduction

Immune responses to SARS-CoV-2 in immunocompromised individuals have been of intense interest throughout the COVID-19 pandemic. In the absence of vaccination, immunologically vulnerable groups are especially susceptible to severe COVID-19 disease and hospitalisation (reviewed in (1)); after vaccination, reduced responses to SARS-CoV-2 vaccines and potential vaccine failure have been of particular concern.

Multiple sclerosis (MS) is an immune-mediated disease characterised by inflammation and demyelination in the central nervous system. Current treatment involves modulation of the immune system to alleviate inflammation. However, some disease-modifying therapies (DMTs) can also impede an effective response to infectious diseases and vaccination (reviewed in (2)). It is unclear whether people with MS (pwMS) are more susceptible to severe COVID-19 disease in the absence of vaccination (3-6); current evidence suggests that this varies depending on DMT usage, where treatment with anti-CD20 drugs presents an increased risk factor (7, 8), as well as neurological disability, comorbidities, and age (6, 8). It is therefore important to establish vaccine effectiveness in pwMS and whether vaccination protects against COVID-19 disease to the same extent as in the general population. Certain DMTs are known to be associated with increased risk of other infections: anti-CD20 drugs such as rituximab, ocrelizumab, and ofatumumab are associated with a range of serious infections, including respiratory tract infections (9, 10); sphingosine-1-phosphate receptor (S1PR) modulators, including fingolimod, ozanimod, and siponimod, which sequester lymphocytes in lymph nodes, are associated with increased risk of herpesvirus infections or reactivations (11); and natalizumab, an anti-alpha-4 integrin monoclonal antibody (mAb), with a risk of progressive multifocal leukoencephalopathy (9).

The primary focus of many vaccine efficacy studies to date has been the humoral immune response (12, 13). Many DMTs, particularly anti-CD20 drugs, target B cells and people on such treatments have reduced or non-existent antibody responses after vaccination (14-18); pwMS on fingolimod have been found to have significantly reduced antibody responses (14, 19). By contrast, pwMS taking other DMTs including natalizumab (20, 21), cladribine (an adenosine mimic which triggers lymphocyte apoptosis) (22), and alemtuzumab (an anti-CD52 mAb that depletes T and B cells) (23) appear to have antibody responses comparable to untreated control groups.

Nevertheless, it is unclear if vaccine-specific T cell responses are impaired. Several studies have looked at cellular responses to SARS-CoV-2 spike peptides in pwMS after two doses of SARS-CoV-2 vaccine (16, 21, 24-26) and found that IFN-γ+ T cell responses were detectable in many, though not all, patients on a variety of DMTs. One exception was pwMS treated with fingolimod (26), where T cell responses were significantly attenuated. Although not all individuals on anti-CD20 therapies developed T cell responses, it further appears that T cell responses and antibody titres are not well correlated, and so a lack of antibody response is not in itself indicative of a failed response to vaccination (27). Additionally, data on the effect of a third vaccine dose on both antibody levels and T cell responses are mixed; some studies suggest no effect of additional vaccination on either humoral or cellular immune responses (28), whereas others find boosted responses (29, 30).

The time period between receiving a dose of DMT and vaccination varies between DMTs. Fingolimod, for example, is taken daily, whereas anti-CD20 treatments are administered at six-month intervals, with a clear impact of this interval on the humoral response. An increased gap between administration of anti-CD20 therapies and vaccination is associated with stronger antibody responses (15, 27, 31, 32), which may be beneficial during vaccination but can also lead to interruptions in ongoing treatment of MS or undesirable delay in vaccine schedules. It is therefore of interest to establish what effect ongoing DMT treatment has on vaccine responsiveness during both the primary vaccine course and for subsequent boosters.

Recent register studies indicate that pwMS treated with high efficacy DMTs, including alemtuzumab, natalizumab, cladribine, S1PR modulators, and anti-CD20 therapies, have the best long-term outcomes for reduced worsening of disability and relapse outcomes (33, 34).

Although for safety reasons alemtuzumab is rarely given to newly diagnosed patients, many people have been treated with this induction therapy during the last decade and comprise an important subset of pwMS. This study therefore focused on the cellular response to these five therapies that are among the most likely to be the treatments of choice for future pwMS.

The aim of this study was to investigate IgG antibody binding to the receptor binding domain (RBD) on the spike protein of SARS-CoV-2 as well as functional spike-specific CD4 and CD8 T cell responses from pwMS on five different DMTs and a healthy control group after two doses of SARS-CoV-2 vaccine. We also investigated whether a third vaccine dose improved the humoral and/or cellular responses in individuals treated with rituximab or fingolimod who had impaired IgG anti-spike RBD antibody responses after two vaccine doses.

## Results

A cohort of pwMS living in Oslo or Akershus, Norway, who were treated with DMTs prior to the beginning of the COVID-19 pandemic were recruited as part of an ongoing population-based study of vaccine responses in pwMS in Norway (NevroVax) (15, 29). Cellular samples were collected from a subset of individuals on different DMTs (fingolimod, rituximab, cladribine, natalizumab and alemtuzumab) both before and after the primary course of 2 vaccine doses in April-July 2021 (Supplementary Figure 1). Healthy controls were recruited from among healthcare workers at Diakonhjemmet Hospital and Akershus University Hospital. Rituximab- and fingolimod-treated individuals who had low antibody responses after vaccination were offered a 3^rd^ vaccine dose in the summer of 2021 (EudraCT Number: 2021-003618-37, see Methods), before recommendations for booster vaccines in Norway were changed in September of that year to recommend a 3^rd^ dose for all immunocompromised individuals. The characteristics of this cohort are described in Table 1 according to DMT, including age, sex, time since last drug administration, and vaccine type (primarily the mRNA vaccines BNT162b2 (Comirnaty, Pfizer-BioNTech) and mRNA-1273 (Spikevax, Moderna); see Methods for further details). Breakthrough COVID-19 infections >14 days after vaccination are shown for each group (n=29 across all DMTs and healthy controls). Infections were predominantly contracted between November 2021-February 2022, representing a mixture of Delta and Omicron VOC infections. None of the individuals were hospitalised or died.

**Table 1:**
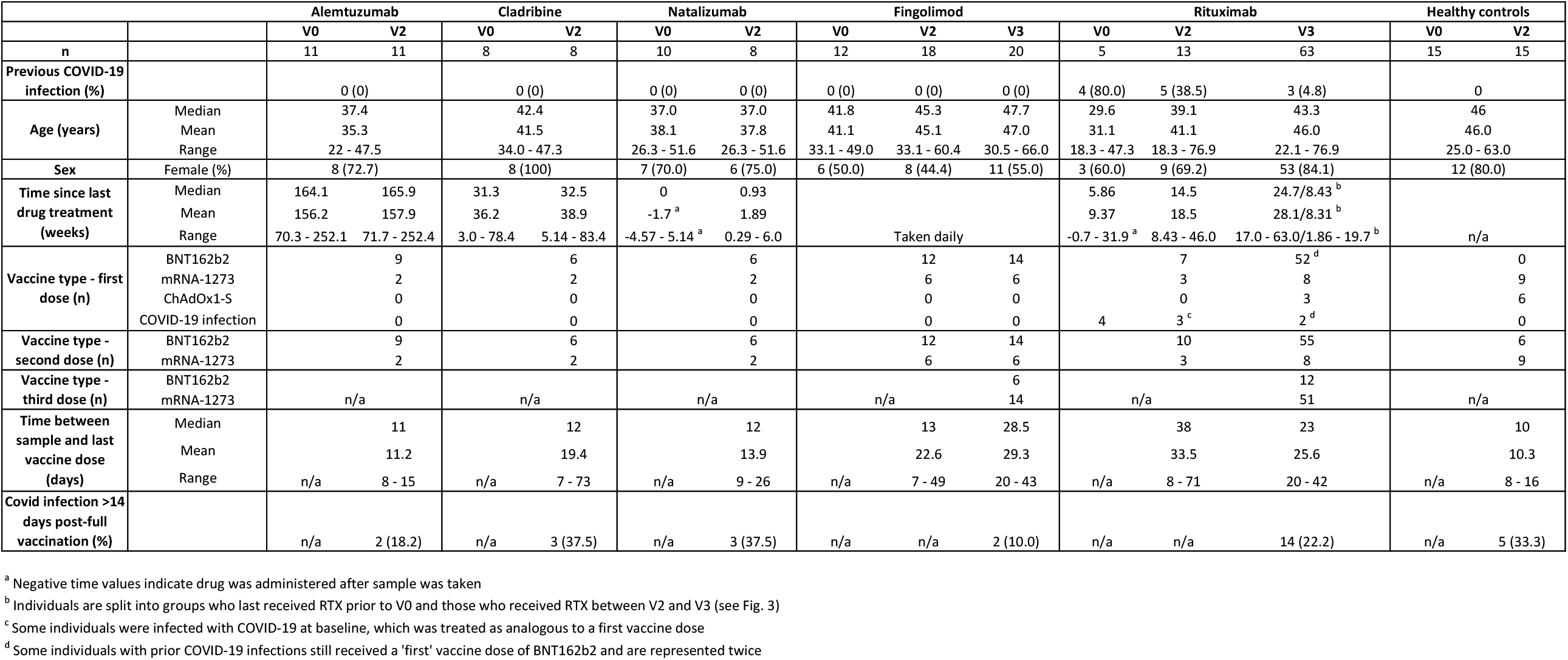
Summary of participant characteristics. Participants are grouped by DMT. V0 indicates pre-vaccination baseline samples, V2 indicates samples taken after receiving 2 doses of vaccine, and V3 indicates samples taken after 3 doses of vaccine. Numbers of individuals per DMT group, age, sex, time since last drug treatment, vaccine types, time between sampling and the corresponding vaccine dose, and the number of participants in each group who were subsequently infected with COVID-19 >2 weeks post-vaccination.

DMTs vary based on mechanism of action and cellular target. We therefore assessed the effect of each DMT on the lymphocyte, CD4 and CD8 T cell frequency and function by flow cytometry (Figure 1). The full flow cytometry gating strategy is shown in Supplementary Figure 2. Compared to healthy controls, pwMS on DMTs did not show altered frequencies of CD3+ lymphocytes except for fingolimod-treated individuals, who had significant reductions in the CD3+ cell populations (Figure 1A) (proportion of CD3+ live lymphocytes in fingolimod-treated: median, 46.9%; IQR, 44.9%; healthy controls: median, 66.8%; IQR, 11%). Additionally, fingolimod-treated individuals had sharply reduced CD4+ T cell populations (fingolimod-treated: median, 19.2%; IQR, 26.0%; healthy controls: median, 63.1%; IQR, 14.1%) and a concomitant increase in CD8+ T cells (Figure 1B and Supplementary Figure 3). The graphs show frequencies from the post-vaccination time point; however, the cell population frequencies for individuals were consistent before and after vaccination. The CD4:CD8 T cell ratio did not correlate with antibody responses in fingolimod-treated individuals, and people with less distorted ratios of T cells did not have improved antibody titres, which were low throughout the group (data not shown).

**Figure 1:**
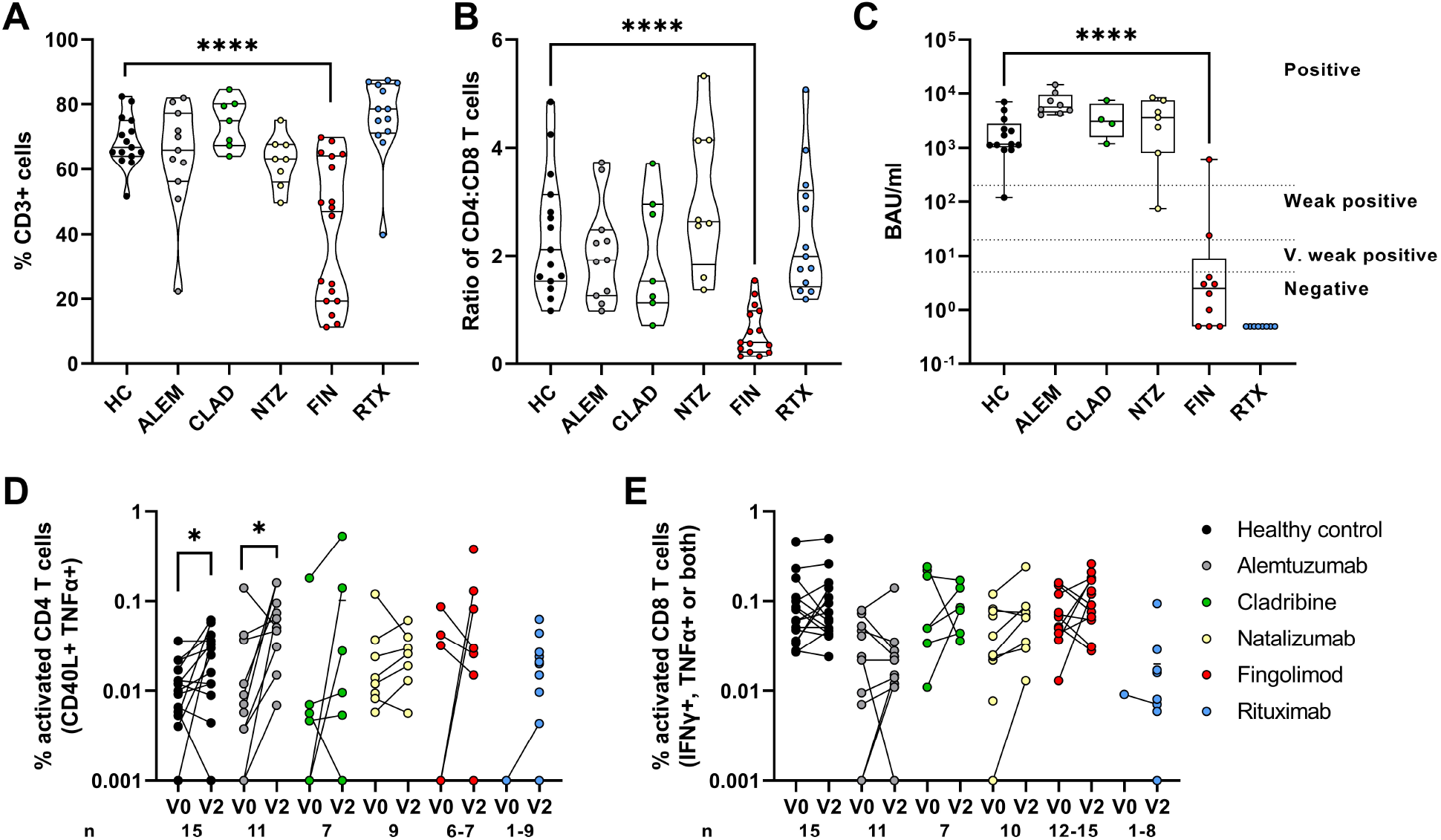
Lymphocyte proportions in peripheral blood and spike-specific vaccination responses in pwMS on DMTs. Individuals are grouped by DMT (healthy controls (HC), alemtuzumab (ALEM)-, cladribine (CLAD)-, natalizumab (NTZ)-, fingolimod (FIN)- and rituximab (RTX)-treated MS patients). (A) CD3+ lymphocyte proportions and (B) the ratio of CD4:CD8 T cells in different DMT groups showed reduced frequencies of CD3+ lymphocytes and CD4+ T cells in fingolimod-treated patients. Violin plots show individuals as separate points, lines indicate median, IQR, and min and max. Mann-Whitney test comparing drug-treated groups with healthy controls, two-tailed p values were calculated, **** p<0.0001. (C) Binding antibody units after 2 doses of vaccine. Responses below the lower limit of detection are shown as 0.5 BAU/ml; titres <5 BAU/ml are considered negative, 5-20 BAU/ml as very weak positives, 20-200 BAU/ml as weak positives and >200 BAU/ml as positives. (D) CD4 T cell (CD40L+ TNF-α+) and (E) CD8 T cell responses (IFN-γ+ and/or TNF-α+) to spike peptides before (V0) and after (V2) 2 doses of vaccine. Responses with 0 events are plotted at 0.001% to indicate non-responses. Samples from the same individual before and after vaccination are paired with a line. Patient numbers for each group are indicated along the x-axis. Individuals with <1000 CD4 or CD8 T cells acquired by FACS were excluded from further analysis. Statistical comparisons by Wilcoxon two-tailed paired t-tests, * p<0.05.

Antibody and T cell responses were measured to assess the immune response to two doses of vaccine. Samples taken 3 weeks (median 20.5 days) after the 2^nd^ vaccine dose were assayed for antibody binding activity (Figure 1C). IgG anti-spike RBD responses were classified as negative (<5 BAU/ml), very weak positive (5-20 BAU/ml), weak positive (20-200 BAU/ml), and positive (>200 BAU/ml) and are indicated on the graph for reference. All healthy controls and individuals treated with alemtuzumab, cladribine and natalizumab had strong antibody titres, predominantly in the ‘positive’ range (median per group: healthy control, 1166 BAU/ml; alemtuzumab, 5591 BAU/ml; cladribine, 3081 BAU/ml; natalizumab, 3625 BAU/ml). However, individuals treated with fingolimod or rituximab had poor antibody responses after two vaccine doses (median: fingolimod, 2.5 BAU/ml; rituximab, 0.5 BAU/ml (below the level of detection for this assay)).

T cell responses were assessed using activation-induced marker (AIM) assays and measured by flow cytometry (see Supplementary Figure. 2 for gating). Peripheral blood mononuclear cells (PBMCs) were stimulated with SARS-CoV-2 spike peptides and CD4 T cell activation was measured by CD40L and TNF-α coexpression (Figure 1D) before (V0, baseline) and 2 weeks after (V2) vaccination. Samples were taken from the same individuals at both time points wherever possible, indicated by paired dots. There was a significant increase in the spike-specific CD4 T cell response after vaccination in the healthy controls and alemtuzumab-treated patients. This suggested that most of the alemtuzumab-treated pwMS had reconstituted their immune system within the time since last treatment, which was more than three years for most patients. Responses were highly heterogeneous and did not reach statistical significance in the other DMT groups. More than half (10/18) of the fingolimod-treated group had too few CD4 T cells in our assay to accurately measure activation responses and were excluded from the analysis as we had too few CD4+ events to calculate the percent response. CD8 T cell responses (Figure 1E) producing IFN-γ and TNF-α varied between individuals. Of interest, IFN-γ+ and TNF-α+ spike-specific CD8+ T cell responses from fingolimod-treated individuals negatively correlated with higher proportions of CD8+ T cells (i.e. individuals with more skewed CD4/CD8 ratios also had fewer cytokine producing CD8 T cells), although this did not reach statistical significance (Supplementary Figure 3I).

As the fingolimod- and rituximab-treated individuals had poor antibody responses, these patients received a third dose of vaccine (see Methods) (Figure 2). Individuals treated with rituximab did not show significant improvements in IgG anti-spike RBD after a third vaccine dose (Figure 2A), although the overall responses and number of responders increased (median and IQR at V2, 1.3 and 18.3 BAU/ml; at V3, 2.0 and 665.5 BAU/ml; 13/43 (30.2%) individuals had >5 BAU/ml titres after 2 vaccine doses, increasing to 27/61 (44.3%) after 3 vaccine doses), suggesting that some, though not all, individuals improved their antibody responses after repeated vaccination. However, the number of patients that were positive (>200 BAU/ml) was significantly increased from 5/43 after 2 doses to 19/61 after 3 doses (two-tailed p=0.0320, Fisher’s exact test). Moreover, a significant proportion of individuals had detectable spike-specific CD4 and CD8 T cell responses (Figure 2B-C), demonstrating that rituximab treatment does not inhibit T cell responses to the same extent as antibody responses.

**Figure 2:**
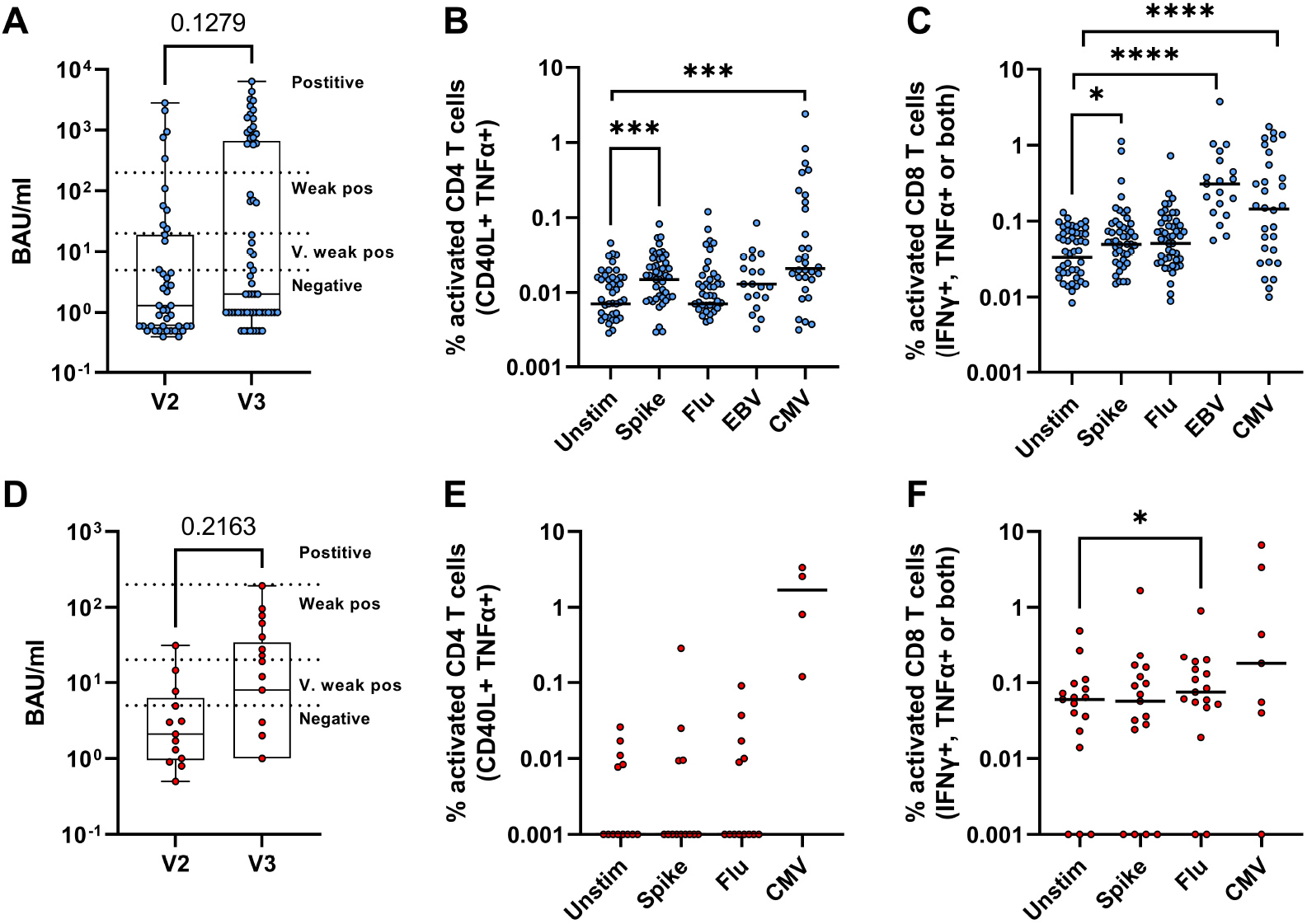
T cell and antibody responses in rituximab- and fingolimod-treated patients after 3^rd^ vaccine dose. Antibody responses (BAU/ml), CD4+ and CD8+ T cell responses after 3^rd^ vaccine dose in rituximab-treated (A-C) and fingolimod-treated patients (D-F) (A, n=43-61); B, n=21-56; C, n=21-54; D, n=13-21; E, n=6-13; F, n=6-17). Individuals with <1000 CD4+ T cells acquired by FACS were excluded from this analysis. Dotted lines in (A) and (D) indicate classification of antibody responses as negative or positive as described previously. For B-C and E-F, lines on scatter plots indicate the median. Statistical analyses by Wilcoxon paired t-tests, two-tailed p values are shown, * p<0.05, *** p<0.001

The same effect was not seen for people treated with fingolimod. After a third vaccine dose, fingolimod-treated patients showed no significant increase in IgG anti-spike RBD (Figure 2D), and generally had even lower antibody responses than the rituximab-treated group, with no patients reaching the ‘positive’ response classification of >200 BAU/ml (median and IQR at V2, 2.1 and 5.35 BAU/ml; at V3, 8.0 and 33.5 BAU/ml). Despite an increase in weak responders (20-200 BAU/ml), this was not statistically significant (1/13 individuals had weak responses after V2 compared to 7/21 after V3; two-tailed p=0.1164, Fisher’s exact test). Spike-specific CD4 and CD8 responses also showed no significant response, suggesting that fingolimod has a major impact on measurable T cell responses in blood as well as antibody levels. We did see a small but statistically significant CD8 T cell response (p=0.0429) to influenza (flu) peptides compared to the unstimulated control, suggesting that existing T cell responses, possibly generated prior to beginning fingolimod treatment, are maintained over time.

61.4% (51/83) of fingolimod- or rituximab-treated patients received an influenza vaccine between September 2020 and February 2021. However, influenza-specific T cell responses did not significantly differ between individuals who had received a seasonal influenza vaccination during the previous winter (2020-21) and those who had not. This suggested that T cell responses generated via previous vaccination or influenza infections prior to the COVID-19 pandemic were still detectable in these patients. There was a weak but significant positive correlation between CD4 responses to SARS-CoV-2 spike and CD4 responses to influenza peptides in rituximab-treated patients, suggesting individual differences to vaccine antigens in general (Supplementary Figure 4A). T cell responses to EBV and CMV peptides were higher than responses to the vaccine peptides, which represents the difference between vaccination and latent viral infection. In rituximab-treated individuals we saw strong CMV-specific CD4 and CD8 T cell responses (p=0.0005 and p<0.0001 respectively, Wilcoxon tests) (Figure 2B) and EBV-specific CD8 T cell responses (p<0.0001) (Figure 2C). However, there was no correlation between CD4 T cell responses to spike and CMV in rituximab-treated patients, (Supplementary Figure 4B), CD4 and CD8 T cell responses to spike (Supplementary Figure 4C), or CD4 responses and antibody responses (Supplementary Figure 4D), consistent with other studies showing low concordance between these measures of immune responsiveness (27).

The administration interval of DMTs varies by drug, as described in Table 1. In the course of this study, patients taking rituximab received treatment according to their individual schedules. All patients received rituximab prior to the baseline (V0) sample and completed the initial two-dose vaccine course without further rituximab infusions. Between the second and third vaccine doses, approximately half (30/62) of the patients received another dose of rituximab (median time 8.43 weeks before V3, range 1.86-19.7 weeks). We hypothesised that this rituximab dosage impaired the ability to respond to vaccination. Antibody and T cell responses in these two groups were therefore compared (Figure 3). There was no significant difference in IgG anti-spike RBD between these two groups after the second vaccine dose (V2), but after a third vaccine dose (V3) individuals who had recently received rituximab had significantly lower antibody activity (Figure 3A) than those who had not (p=0.023, unpaired t test). However, there was no such difference between the T cell responses of the two groups (Figure 3B-C). Comparing the spike-specific responses between groups showed no difference in CD4 (p=0.998, unpaired t test) or CD8 T cell activation (p=0.545), suggesting that T cell responses are not affected by re-administration of rituximab after the primary vaccine course.

**Figure 3:**
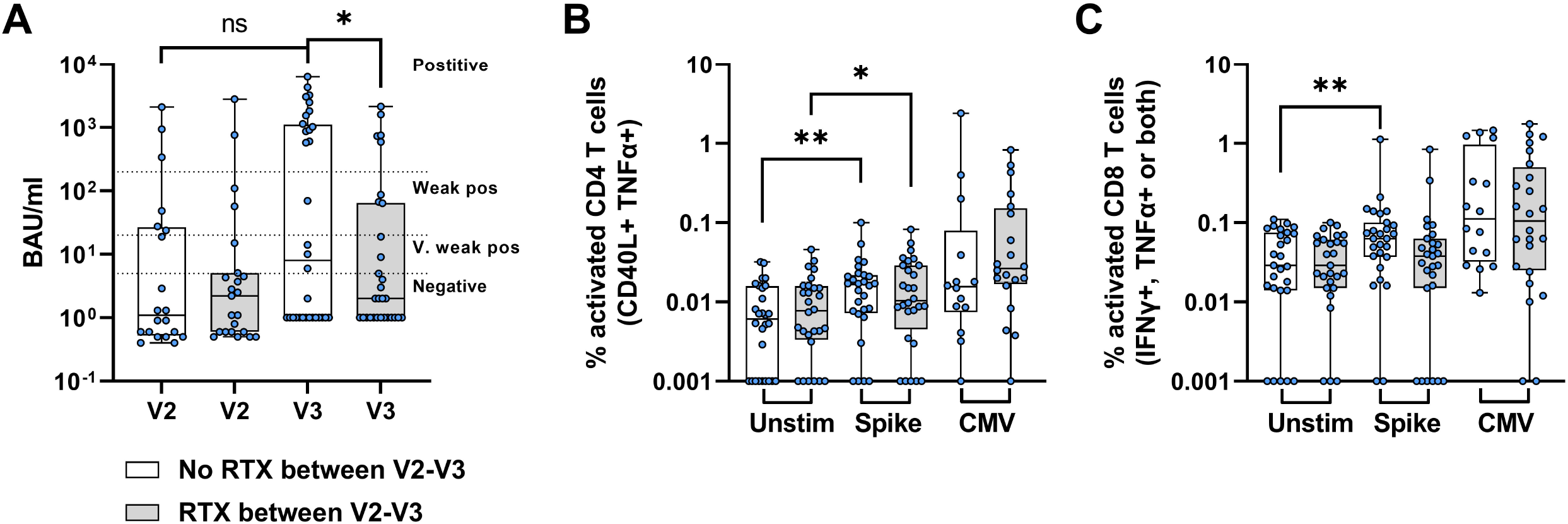
Re-administration of rituximab between vaccine doses affects antibody but not T cell responses. Rituximab-treated individuals were grouped by whether or not they received a dose of RTX between vaccine doses 2 and 3. (A) Antibody titres (BAU/ml) after 2^nd^ (V2) and 3^rd^ (V3) vaccine doses for patients who did not receive RTX between vaccines (empty boxes) (n= 20-32) and patients who did receive RTX between vaccines (grey boxes) (n=23-30). (B) CD4 T cell and (C) CD8 T cell responses without stimulation (unstim) or to SARS-CoV-2 spike or CMV peptides after 3^rd^ vaccine dose for patients without (n=28) or with RTX administration (n=28) between vaccine doses, as previously. Statistical analyses for paired responses by Wilcoxon t test, unpaired responses by Mann-Whitney, two-tailed p values are shown, * p<0.05, ** p<0.01.

Finally, the question of whether vaccination confers protection against SARS-CoV-2 variants of concern (VOC) has been of particular concern since the initial emergence of the Alpha (B.1.1.7) variant and subsequent Delta (B.1.617.2) and Omicron (B.1.1.529/BA.1-5) variants. Mutations in the spike region of these variants are thought to reduce the ability of vaccine-generated antibodies to recognise these variants and potentially to reduce protection against them. To measure how T cell responses from vaccination were affected, we assessed CD4 and CD8 T cell responses to the mutated peptides of these three variants (Figure 4). PBMCs from triple-vaccinated rituximab-treated patients were stimulated as before with only the mutated peptide regions from the Alpha, Delta, or Omicron variants, as well as the homologous peptides for each variant from the original Wuhan-Hu-1 sequence). The location and number of mutated peptides (34, 32 and 83 peptides for Alpha, Delta, and Omicron respectively) are shown in Figure 4A. There were no significant differences in CD4 T cell responses to the Alpha variant compared to the homologous Wuhan-Hu-1 sequence, but significantly reduced responsiveness to the mutated peptides of both the Delta (p=0.047, Wilcoxon t test) and Omicron variants (p=0.0028) (Figure 4B). Although CD8 T cell responses were reduced, particularly for the Delta VOC, these differences did not reach significance (Figure 4C).

**Figure 4:**
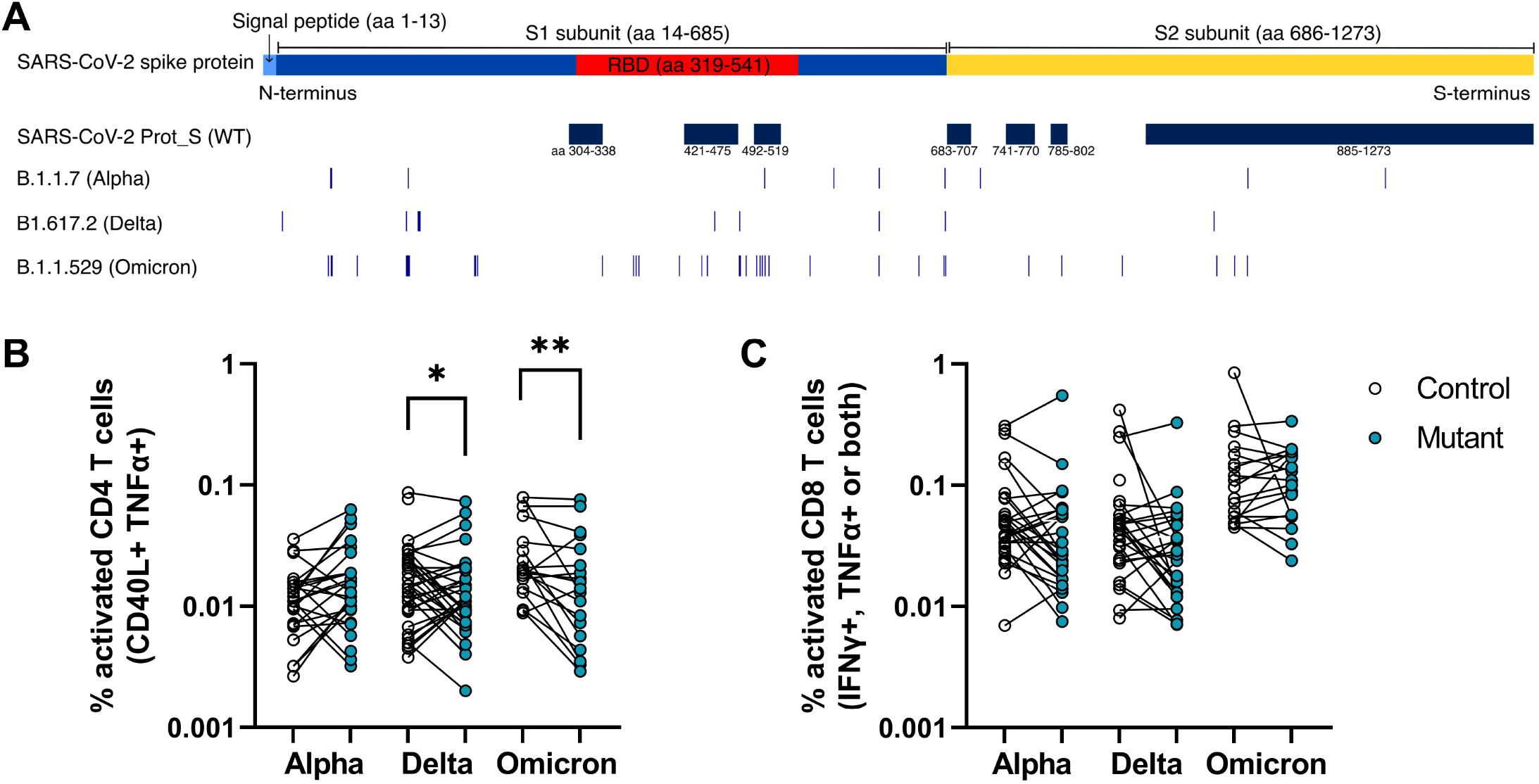
T cell responses to the Delta and Omicron SARS-CoV-2 variants after three vaccine doses. Schematic of mutated regions in the Alpha, Delta and Omicron regions stimulated by peptides (A). The SARS-CoV-2 spike protein is 1273 amino acids (aa) long, consisting of the signal peptide, S1 and S2 subunits; the receptor binding domain (RBD) in S1 is indicated in red (54). Regions covered by the SARS-CoV-2 Prot_S (WT) peptide used for AIM assays are shown for reference. The control and mutant peptides for each variant cover the same loci but with the mutated (mutant) or Wuhan-Hu-1 variant (control). Amino acid mutations are listed in Methods. PBMCs from rituximab-treated patients after a 3^rd^ vaccine dose were stimulated with spike peptide pools from the mutated regions (blue circles) of the Alpha (n=29), Delta (n=41) and Omicron VOC (n=21) and the same regions of the WT sequence (empty circles) and the CD4+ (B) and CD8+ (C) T cell responses were compared. Statistical differences were calculated by Wilcoxon paired t tests. * p<0.05, ** p<0.01

## Discussion

Older and immunocompromised individuals are particularly at risk of severe COVID-19 disease. Vaccine efficacy in immunocompromised individuals is therefore important to understand, particularly as many countries including Norway (35) have achieved high vaccine coverage and have since lifted many or all infection-limiting measures such as social distancing. However, SARS-CoV-2 variants continue to circulate and vulnerable groups may still be at risk of severe disease. These data show that pwMS treated with alemtuzumab, cladribine, and natalizumab have robust humoral and CD4 and CD8 T cell responses after two vaccine doses, in agreement with other studies (3, 14, 20, 22, 25). However, individuals treated with fingolimod and rituximab have strongly reduced antibody responses compared with both healthy controls and pwMS taking other DMTs. Upon receipt of a third vaccine dose, both treatment groups showed small increases in IgG anti-spike levels and a significantly increased percentage of patients developed high responses (>200 BAU/ml) in the rituximab treated group, demonstrating that some individuals were capable of increasing B cell responses. This finding was also found in a larger study where improved IgG anti-spike responses were found after third vaccination (29). Moreover, triple-vaccinated rituximab-treated individuals demonstrated both CD4 and CD8 T cell responses against SARS-CoV-2 spike peptides. These T cell responses were not reduced even when individuals received rituximab between their second and third vaccine doses, suggesting that although re-administration of anti-CD20 drugs does impair humoral responses, cellular responses are preserved.

Additionally, we observed strong CD4 and CD8 T cell responses to the herpesviruses CMV and EBV in pwMS, suggesting that specific T cell responses against antigens from long-term latent infections are present. T cell activation against other vaccine antigens such as influenza were comparable to the SARS-CoV-2 spike-specific responses, and rituximab-treated individuals showed a positive correlation between spike-specific and flu-specific responses. This suggests that vaccine responsiveness varies by individual but is not necessarily associated with T cell responses to other infections such as CMV. Immune responses may also differ based on whether individuals were already using DMTs at the time of antigen exposure, which may affect the magnitude of the immune response, as well as the availability and duration of antigen seen during vaccination or acute infection compared to chronic infections.

In pwMS treated with fingolimod, a third vaccine dose did not appear to improve either the antibody or T cell responses. As fingolimod is taken daily, the fluctuations in B cell counts seen in individuals taking anti-CD20 drugs are not seen (36). The reduction in peripheral lymphocyte and CD4 T cell counts we observed here was consistent in individuals at different sampling points, suggesting that the administration of fingolimod causes lymphocyte sequestration to different extents for each individual. Although antibody titres were strongly reduced for all fingolimod-treated patients, we observed that individuals with less skewed CD4:CD8 T cell ratios had stronger spike-specific CD8 T cell responses, suggesting that people with higher circulating CD4 T cell frequencies are more likely to generate measurable and potentially protective cellular responses.

Nevertheless, pwMS receiving fingolimod do not appear to be at higher risk of severe COVID-19 or hospitalisation than the general population prior to vaccination (7, 37). Fingolimod has been found to reduce proinflammatory cytokine release from dendritic cells and monocytes (38) which may reduce detrimental uncontrolled inflammation associated with severe COVID-19 disease (39). Additionally, as lymphocytes are sequestered rather than destroyed by S1PR modulators (40), failure to detect T cell responses in peripheral blood may not fully reflect the extent of the total T cell response, and non-circulating cellular responses induced by vaccination may be present in the lymph nodes or other secondary lymphoid organs.

Several large-scale studies have found that pwMS on fingolimod or ocrelizumab are at higher risk of SARS-CoV-2 infection after vaccination than the general population or pwMS on other DMTs (41-43), possibly reflecting the role of circulating antibodies in preventing infection. However, the severity of these infections is still unclear, and where cases could be followed on an individual level, there were no deaths from COVID-19 (42, 43). Further research has found that, even after vaccination, pwMS on anti-CD20 drugs were at higher risk of hospitalisation but not death; this risk was not seen with other DMTs including S1PR modulators (44). In our study, 29 individuals across all DMTs contracted SARS-CoV-2 after vaccination and none of these were hospitalised or died. Further large-scale studies are required to determine vaccine protection against severe disease and death in pwMS and particularly patients treated with anti-CD20 drugs and S1PR modulators.

The emergence of SARS-CoV-2 VOC has further complicated the question of vaccine efficacy and protection. Neutralising antibodies against the Delta and Omicron VOC have been found to be sharply reduced compared to the original Wuhan-Hu-1 strain (45), while T cell responses are more heterogeneous and show wide cross-reactivity to other human coronaviruses as well as between variants (46-48). In triple-vaccinated rituximab-treated individuals we found that CD4+ T cell responses to both the Delta and Omicron VOC were reduced compared to WT, suggesting that although T cells are responsive to the mutated VOC regions, vaccine-generated T cell-mediated protection may be reduced. However, these mutated regions cover only a fraction of the spike peptide sequences and further work is needed to determine how these mutations affect T cell vaccine responses.

We found no correlation between any combinations of antibody titre, CD4 T cell responses, or CD8 T cell responses, and therefore using only one of these parameters as an indication of immune responsiveness cannot give a full picture of vaccine efficacy. Although the correlates of protection against SARS-CoV-2 infection and severe COVID-19 disease are still unclear and the relative roles of antibody-mediated virus neutralisation and T cell-dependent protection are still being extensively studied (49-51), analysis of cellular responses in addition to antibody titres can give a better understanding of whether immunosuppressed individuals are likely to require additional protective measures. Further follow up studies are required to determine whether T cell responses in the absence of antibody titres, such as seen in our rituximab-treated population, are protective against severe disease, but the current evidence supports the contention that T cell immunity is sufficient.

One limitation of this work is a lack of longitudinal sampling to measure changes in CD4 and CD8 T cell responses between the second and third vaccine for the rituximab- and fingolimod-treated patients. The question of whether repeated vaccination with antigens from the Wuhan-Hu-1 variant can prevent disease from successive VOC remains to be seen.

In summary, we found that pwMS on DMTs that inhibit antibody responses are still capable of mounting T cell responses comparable with healthy controls, and furthermore that continued administration of the widely used anti-CD20 drug rituximab between the primary vaccine course and subsequent vaccine doses does not impede cellular responses. Further analyses of the efficacy and durability of cellular responses, and well as the impact of additional vaccination, are needed to better understand how vaccines protect against severe disease in immunocompromised individuals.

## Methods

### Participant recruitment and ethical approvals

All patients from the Norwegian MS registry (n=12000) in 2021 were invited to participate in the humoral arm of the NevroVax study. A subgroup of patients from Oslo University Hospital on the DMTs alemtuzumab, cladribine, natalizumab, fingolimod and rituximab (c. n=10 per DMT) were recruited to provide PBMC samples, along with all patients who lacked antibody responses after 2 vaccine doses (considered at the time to be <70 arbitrary units (AU)/ml by ELISA). Individuals from Oslo University Hospital, Akershus University Hospital and Haukeland University hospital with low humoral responses subsequently received a third vaccine dose and those treated at Oslo University Hospital or Akershus University Hospital comprised the fingolimod- and rituximab-treated individuals at V3. Healthy controls were recruited among healthcare workers from Diakonhjemmet Hospital and Akershus University Hospital and samples were stored in the Oslo University Hospital biobank.

### Vaccination and inclusion in vaccination trial

PwMS were vaccinated as per guidelines of the Norwegian Corona Vaccination Program where immunocompromised individuals and healthcare workers (who participated here as healthy controls) were high priority. Vaccines were administered according to the manufacturers’ recommendations and health administration advice at the time, ranging from three weeks between first and second doses for mRNA-1273 and 6-10 weeks for BNT162b2. Some individuals received first doses of ChAdOx1-S (Vaxzevria, AstraZeneca), the distribution of which was subsequently discontinued in Norway in March 2021, and received second doses of BNT162b2. Individuals who had a COVID-19 infection before or during the course of vaccination were excluded from further analyses. Individuals who failed to seroconvert to IgG anti-spike (RBD) after the standard two doses were invited to participate in a vaccination trial to receive a third dose of mRNA-1273 or BNT162b2 outside the framework of the Norwegian Corona Vaccination Program (EudraCT Number: 2021-003618-37). Further patients included in this study after 1^st^ September 2021 received third dose vaccines following revised guidelines in the Norwegian Corona Vaccination Program (where all immunocompromised adults were advised to receive a third dose).

### Sample collection

Venous blood for PBMC isolation was collected at Oslo University Hospital into BD Vacutainer CPT tubes with sodium citrate. Tubes were centrifuged for 20 minutes at 1600xg to isolate PBMCs, which were then pipetted into fresh tubes, washed twice with RPMI, and frozen in 90% foetal calf serum (FCS) (Gibco, Thermo Fisher) with 10% dimethyl sulfoxide (DMSO, Sigma Aldrich) in liquid nitrogen for future use.

### T cell stimulation and flow cytometry

Cryopreserved PBMCs were thawed in RPMI, washed thrice to remove residual DMSO, and counted. Cells were plated into 96-well U-bottomed plates at 200,000 cells per well and stimulated for 24 hours in RPMI culture media containing 10% FCS, 1mM sodium pyruvate (Gibco, Thermo Fisher), 1x MEM NEAA (Gibco), 50nM 1-thioglycerol and 12ug/ml gensumycin. GolgiPlug (BD Biosciences) containing brefeldin A was added after 2 hours of stimulation until the end of the incubation. Cells were stimulated with peptide pools: PepTivator SARS-CoV-2 Prot_S covering the immunodominant sequence domains of the spike glycoprotein from the SARS-CoV-2 (Wuhan-Hu-1 variant), EBV Consensus, and CMV pp65 pool (used according to the manufacturer’s recommendations at 0.75nmol/ml, all Miltenyi Biotec) and pooled pan-influenza peptides for HLA class I and II (final concentration 1µg/ml) (GenScript). Peptide pools for mutated SARS CoV-2 Spike are outlined in the next paragraph. Cytostim (Miltenyi Biotec) was used as a positive control according to the manufacturer’s recommendation.

After 24 hours, cells were centrifuged at 500xg for 5 minutes, the supernatant discarded, and cells resuspended in FACS buffer (1% FCS in PBS). Cells were centrifuged again and the supernatant removed. Cells were incubated with 10µl surface antibody cocktail (anti-human CD3-BV605 (clone SK7) (BD Biosciences), CD4-eFluor 450 (OKT-4), CD8-AF488 (OKT-8), and Fixable Live/Dead Near-IR (1:1000 dilution) (all ThermoFisher)) for 30 minutes at 4°C, washed in FACS buffer, then fixed in Fix/Perm (BD Biosciences) for 20 minutes at room temperature. Cells were then washed twice with PermWash (BD Biosciences) and incubated with 10µl intracellular antibody cocktail (anti-human IFN-γ-BV711 (clone 4S.B3), CD40L-BV510 (24-31) (both BioLegend), TNF-α-PE (Mab11), CD69-APC (FN50) (both BD Biosciences)) for 30 minutes at room temperature. Cells were finally washed with PermWash and resuspended in 200ul FACS buffer for analysis by flow cytometry within 24 hours.

Cells were acquired on a BioRad ZE5 flow cytometer and analysed with FlowJo™ v.10.7 Software (BD Life Sciences).

### Variants of Concern and mutated peptide sequences

Three PepTivator SARS-CoV-2 VOC spike protein Mutation Pools and the three corresponding spike protein WT Reference Pools (all Miltenyi Biotec) were used at a final concentration of 0.75 nmol/ml per the manufacturer’s recommendation. Prot_S B.1.1.7 Mutation Pool (cat. no. 130-127-844) included 34 peptides from 10 mutations: deletion 69, deletion 70, deletion 144, N501Y, A570D, D614G, P681H, T716I, S982A, D1118H. The corresponding non-mutated peptide pool control was Prot_S B.1.1.7 WT Reference Pool (cat. no. 130-127-841).

Prot_S B.1.617.2 Mutation Pool (cat. no. 130-128-763) included 32 peptides from 10 mutations: T19R, G142D, E156G, deletion 157, deletion 158, L452R, T478K, D614G, P681R, and D950N. This subvariant lacks the E484Q mutation. The non-mutated peptide pool control was Prot_S B.1.617.2 WT Reference Pool (cat. no. 130-128-761).

Prot_S B.1.1.529/BA.1 Mutation Pool (cat. no. 130-129-928) included 83 peptides from 37 mutations: A67V, H69 deletion, V70 deletion, T95I, G142D, V143 deletion, Y144 deletion, Y145 deletion, N211 deletion, L212I, insertion 214EPE, G339D, S371L, S373P, S375F, K417N, N440K, G446S, S477N, T478K, E484A, Q493R, G496S, Q498R, N501Y, Y505H, T547K, D614G, H655Y, N679K, P681H, N764K, D796Y, N856K, Q954H, N969K, L981F. The non-mutated peptide pool control was Prot_S B.1.1.529/BA.1 WT Reference Pool (cat. no. 130-129-927).

### Antibody quantification

Semiquantitative measurement of antibodies to full-length spike protein (Spike-FL) and the receptor-binding domain (RBD) from SARS-CoV-2 was performed using a multiplexed bead-based assay as described in (52). Polymer beads with fluorescent barcodes were coupled to successively to neutravidin (ThermoFisher) and biotinylated viral antigens to generate bead-based protein arrays. Sera were diluted 1:100 in assay buffer (PBS, 1% Tween-20, 10ug/ml D-biotin, 10 µg/ml neutravidin, 0.1% sodium azide). Diluted sera were incubated with bead-based arrays in 384 well plates for 30 minutes at 22°C at constant agitation, washed three times in PBS/1% Tween-20 (PBT) and labelled with R-Phycoerythrin (R-PE)-conjugated goat-anti-human IgG (Jackson Immunoresearch). For measurement of neutralizing antibodies, the beads were pelleted after incubation with serum and labelled successively with digoxigenin-conjugated human ACE2 and mouse monoclonal anti-dixogigenin (Jackson Immunoresearch), which was conjugated in-house to R-PE. The beads were analyzed with an AttuneNxT flow cytometer (ThermoFisher), and raw data (fcs.3.1) were analyzed in WinList 3D (Verity Softwarehouse). The median R-PE fluorescence intensity (MFI) of each bead subset was exported to Excel. The MFI of beads coupled with viral antigens was divided by that measured on beads coupled with neutravidin only (relative MFI, rMFI). A total of 979 pre-pandemic sera and 810 sera from COVID-19 convalescents were analyzed to establish cutoffs for seropositivity. A double cutoff of rMFI >5 for anti-RBD and anti-Spike FL yielded a specificity of 99.7% and a sensitivity of 95% (53). Serum from an individual who had received three doses of the Pfizer/BioNTech anti-COVID-19 vaccine was used as standard to convert signals to binding antibody units per milliliter (BAU/ml).

### Statistics and analysis

Statistical analyses were performed using GraphPad Prism v.9 for Windows, GraphPad Software. Two-tailed p values are shown. For analysis of functional markers (CD40L+ TNF-α+ CD4 T cells and IFN-γ+ TNF-α+ CD8 T cells), data from FACS plots with fewer than 1000 CD4 or CD8 T cells were excluded.

### Study approval

The study was approved by the Norwegian South-Eastern Regional Ethical Committee (Reference numbers 200631, 235424, 135924, and 204104), and the Norwegian Medicines Agency (EudraCT Number: 2021-003618-37). All participants gave written informed consent prior to inclusion in this study.

## Data Availability

All data produced in the present study are available upon reasonable request to the authors

## Author contributions

LAM and SM conceived and designed the study. A-SW drafted the paper. AR, MK, LAM, TH, GON, and SM contributed to drafting the paper. A-SW, AR, GS, IFK, and AC performed experiments. A-SW, AR, MK, GS, IFK, FL-J, GON, LAM, and SM analyzed and interpreted data. MHØ, AC, TH, HFH, EAH, JTV, and EGC contributed to data analysis and interpretation. MK, GON, MHØ, SWS, KKJ, JTV, FL-J, and LAM organized the collection of samples and information from the cohorts. A-SW, AR and MK are joint first authors with the order determined by contribution to experimental analysis and writing of the manuscript. All authors contributed to and approved the final manuscript.

## Acknowledgements

The authors would like to thank Ingrid Egner, Katrine Persgård Lund, Viktoriia Chaban, and Julie Røkke Osen at the Department of Immunology, Oslo University Hospital for biobanking and technical help. We would also like to thank the participants in the NevroVax study and staff at Diakonhjemmet Hospital and Akershus University Hospital for donating blood samples, as well as the staff at Oslo University Hospital for taking the samples. This study was supported by the Norwegian Institute of Public Health, the Norwegian Ministry of Health through a program for corona vaccination surveillance, the Odd Fellows Foundation, the Sanofi Research Fund for MS, the Norwegian MS Society, and the Research Council of Norway (RCN) Covid (312693), LAM, SM; a KG Jebsen Foundation (grant 19), LAM; the Coalition for Epidemic Preparedness Innovations (CEPI), LAM, SM, JTV, FL-J; and the University of Oslo and Oslo University Hospital, LAM, FL-J, JTV.

