## Supplementary data Figures 1-4 for "T cell responses to SARS-CoV-2 vaccination in people with multiple sclerosis differ between disease-modifying therapies"

Supplementary Figure 1: NevroVax study participants. The primary NevroVax study included humoral responses from 5616 individuals in Norway. A subset of individuals in the Oslo and Akershus areas gave cellular samples for the cellular arm of the NevroVax as described in this paper. The number of individuals in each DMT group and sample time point (V0, V2, V3) are shown here.

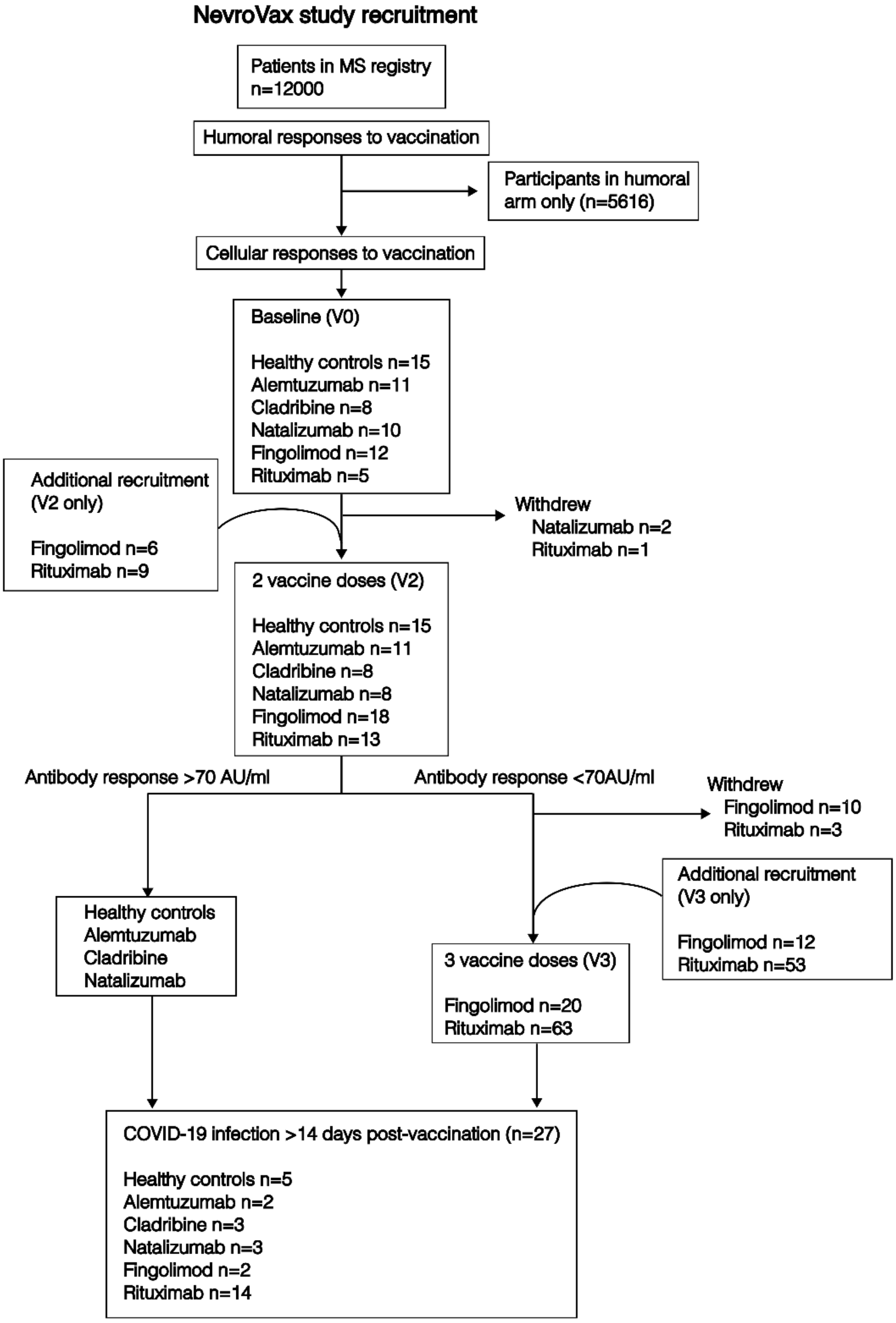

Supplementary Figure 2: Gating strategy for functional CD4 and CD8 responses. Cells were acquired on a BioRad ZE5 flow cytometer and gated for singlets, lymphocytes, live cells, CD3+ lymphocytes, and CD4+ or CD8+ T cells. CD4+ T cells were then gated for CD40L+ TNFα+ double positive events. CD8+ T cells were gated for IFNγ+, TNFα+ or double positive events. Positive events are shown for CMV-stimulated cells.

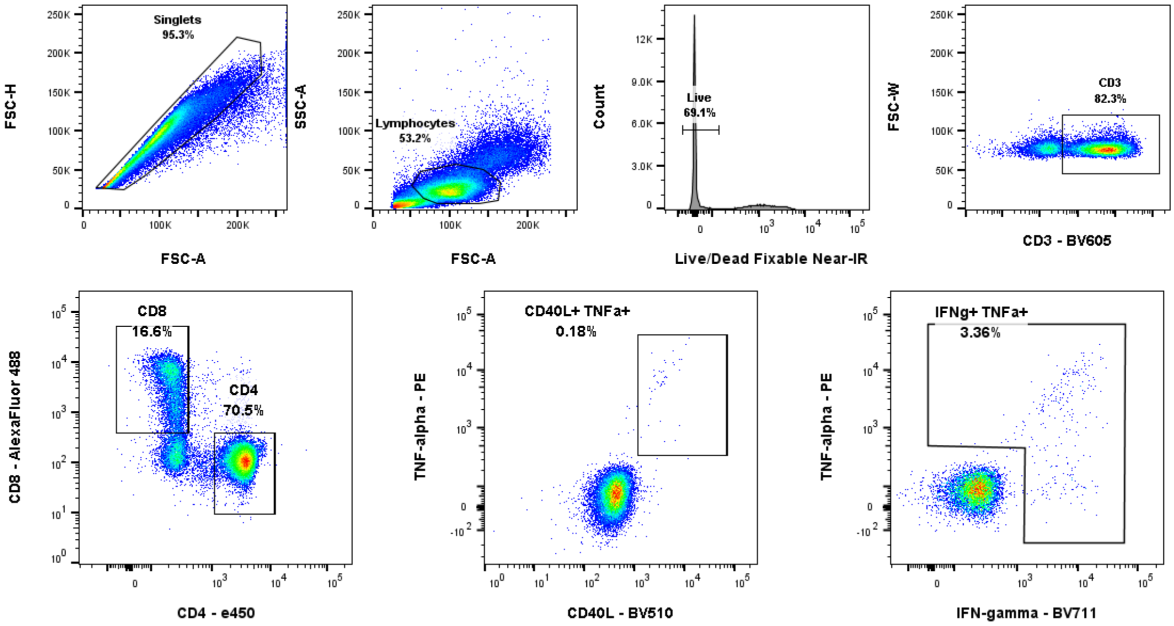

Supplementary Figure 3: CD4+ T cell and CD8+ T cell frequencies are altered in fingolimod-treated pwMS. Example FACS plots showing CD3+ lymphocytes and CD3+CD4+ T cells and CD3+CD8+ T cells from a healthy control (A-B), a less skewed fingolimod-treated individual (C-D) and a more skewed fingolimod-treated individual (E-F). Fingolimod-treated patients have reduced frequencies of CD4 T cells (G) and proportionally increased frequencies of CD8 T cells (H). Statistical comparisons calculated by Mann-Whitney tests, **** p< 0.0001, *** p< 0.001. Fingolimod-treated patients with more skewed CD8+ T cells have a trend towards lower CD8 T cell responses (n=15). Correlation tested by Spearman regression analysis, not significant.

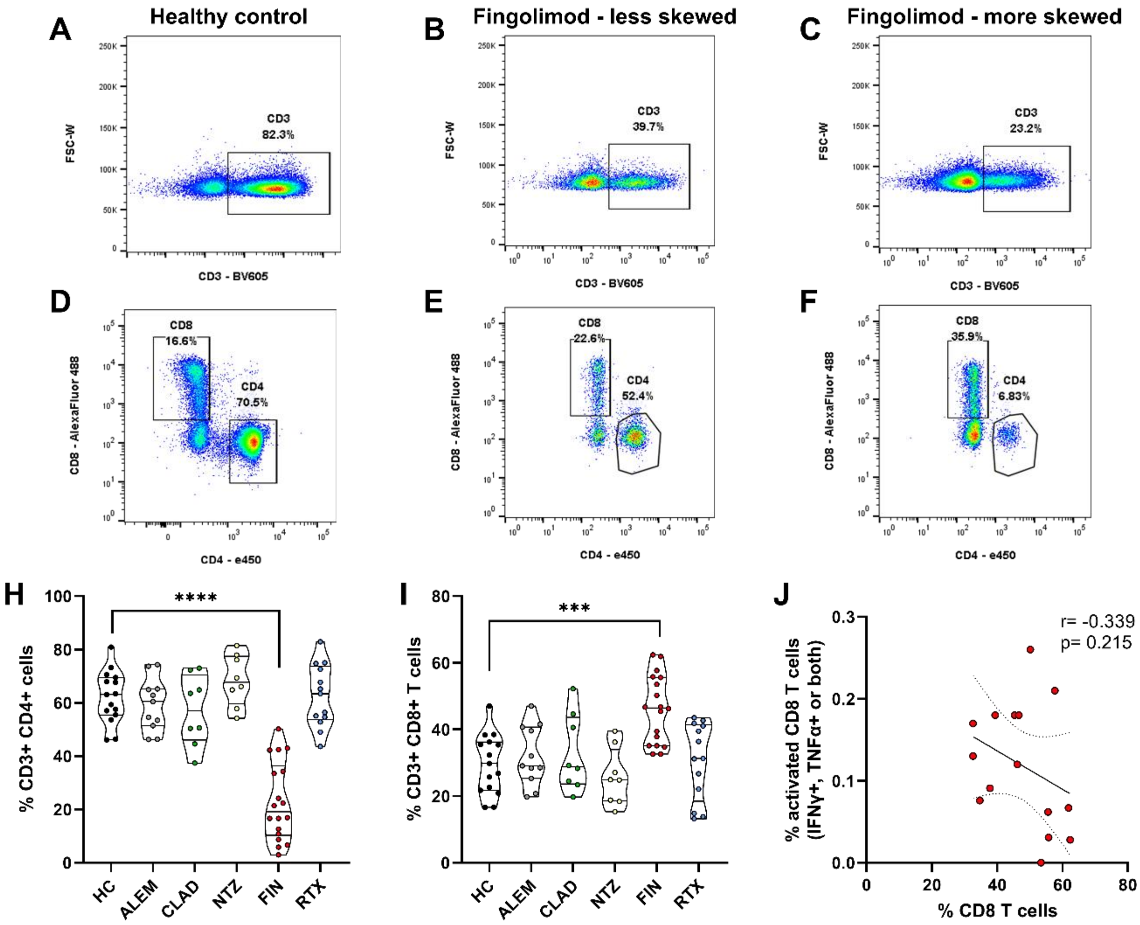

Supplementary Figure 4: Spike-specific CD4 T cell responses to vaccination correlate with influenza-specific responses. Correlations of spike-specific CD4 T cell responses in rituximab-treated individuals after 3rd vaccine dose with (A) CD4 responses to influenza peptides (n=56), (B) CD4 responses to CMV peptides (n=33), (C) CD8 responses to SARS-CoV-2 spike (n=56), and (D) anti-spike antibody titres (BAU/ml) above the negative threshold of 5 BAU/ml (n=27). Linear regression lines are plotted on graphs and r values are indicated on each plot.

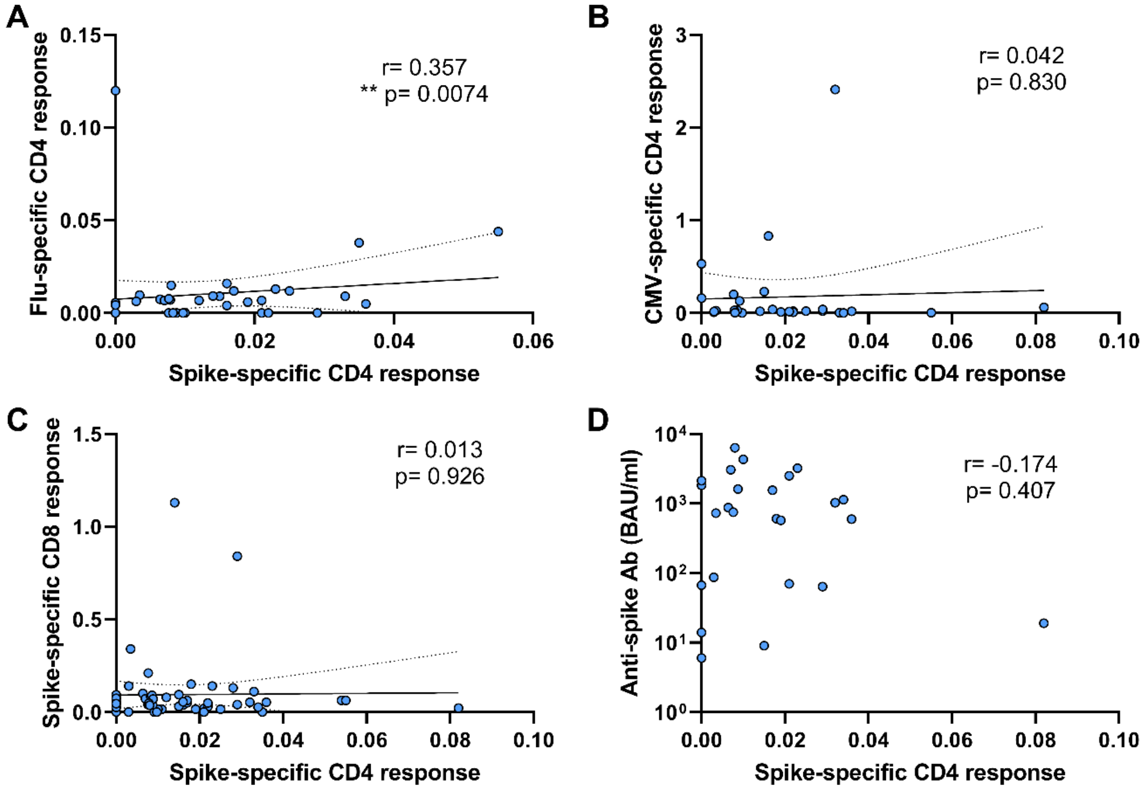
